# Can Vaccine Prioritization Reduce Disparities in Covid-19 Burden for Historically Marginalized Populations?

**DOI:** 10.1101/2021.07.27.21261210

**Authors:** Erik Rosenstrom, Jessica Mele, Julie Ivy, Maria Mayorga, Mehul Patel, Kristen Hassmiller Lich, Karl Johnson, Paul Delamater, Pinar Keskinocak, Ross Boyce, Raymond Smith, Julie L. Swann

**Author notes:** Correspondence: Julie Swann; NC State University; 915 Partners Way; Campus Box 7906, Raleigh, NC 27695.

## Abstract

1.

**Importance:** Nationally stated goals for distributing SARS-CoV-2 vaccines included to reduce COVID-19 mortality, morbidity, and inequity using prioritization groups. However, the impact of these prioritization strategies is not well understood, particularly their effect on health inequity in COVID-19 burden for historically marginalized racial and ethnic populations.

**Objective:** To assess the impact of vaccination prioritization and operational strategies on disparities in COVID-19 burden among historically marginalized populations, and on mortality and morbidity by race and ethnicity.

**Design:** We use an agent-based simulation model of North Carolina to project SARS-CoV-2 infections and COVID-19-associated deaths (mortality), hospitalizations (morbidity), and cases over 18 months (7/1/2020-12/31/2021) with vaccine distribution beginning 12/13/2020 to frontline medical and people 75+, assuming initial uptake similar to influenza vaccine. We study two-stage subsequent prioritization including essential workers (“essential”), adults 65+ (“age”), adults with high-risk health conditions, HMPs, or people in low income tracts, with eligibility for the general population in the third stage. For age-essential and essential-age strategies, we also simulated maximal uptake (100% for HMP or 100% for everyone), and we allowed for distribution to susceptible-only people.

**Results:** Prioritizing Age then Essential had the largest impact on mortality (2.5% reduction from no prioritization); Essential then Age had the lowest morbidity and reduced infections (4.2% further than Age-Essential) without significantly impacting mortality. Under each prioritization scenario, the age-adjusted mortality burden for HMPs is higher (e.g., 33.3-34.1% higher for the Black population, 13.3%-17.0% for the Hispanic population) compared to the White population, and the gap grew under some prioritizations. In the Age-Essential strategy, the burden on HMPs decreases only when uptake is increased to 100% in HMPs. However, the Black population still had the highest mortality rate even with the Susceptible-Only distribution.

**Conclusions and Relevance:** Simulation results show that prioritization strategies have differential impact on mortality, morbidity, and disparities overall and by race and ethnicity. If prioritization schemes were not paired with increased uptake in HMPs, disparities did not improve and could worsen. Although equity was one of the tenets of vaccine distribution, the vaccination strategies publicly outlined are insufficient to remove and may exacerbate disparities between racial and ethnic groups, thus targeted strategies are needed for the future.

## 2. INTRODUCTION

Throughout the COVID-19 pandemic, historically marginalized communities of color have experienced a disproportionate burden of morbidity and mortality^1-7^. The unequal impact on these populations is driven by risk factors such as essential-worker status, age, living arrangements, and high-risk medical conditions^1, 2,4-6^. In response, the National Academies of Sciences, Engineering, and Medicine^7^ (NASEM) drafted a framework for the equitable allocation of SARS-CoV-2 vaccines, which aimed at overall reductions in morbidity and mortality and explicitly outlined approaches to mitigate structural inequities. Despite this guidance, there was substantial variability in vaccine rollout strategies at the state and local levels. All states included health care workers and long-term care facility residents in their initial priority groups, but subsequent phases included prioritization for varying combinations and orderings of groups such as frontline essential workers, educators, congregate living facility residents and staff, those with high-risk medical conditions, and older populations^8^. Which strategies best achieved the goals set out by the NASEM or what approaches will most effectively accomplish future public health targets aiming to reduce disparities remains unknown.

Vaccination prioritization scenarios have been studied using various modeling approaches^9-14^. The findings suggested that prioritizing older adults had the greatest impact on COVID-19 mortality, while prioritizing highly interactive individuals had the greatest impact in reducing morbidity (e.g., incidence of disease)^9-12^. Fujimoto et al.^14^ studied susceptible-only distribution (i.e., distributing vaccines only to people without a previous case of COVID-19) and found it to be effective for increasing the benefit of COVID-19 vaccine supply. Ferranna et al.^11^ assessed equity by assuming essential workers in their model are more likely to be members of vulnerable populations. They reported that prioritizing older adults over vulnerable populations led to higher reduction in mortality; however, they did not explicitly consider racial and ethnic disparities that may arise under different vaccination scenarios. To our knowledge, no studies have explicitly designed or tested vaccination strategies explicitly considering race and ethnicity in the models, nor tested additional vaccination strategies aimed at increasing uptake within population subgroups.

Vaccine equity discussions and efforts focused on reducing health inequities that are related to systemic social injustices, e.g., targeting those at disproportionate risk for COVID-19^15^ through measures such as identifying zip codes based on incidence rates or disadvantage indices^16^ (e.g., social vulnerability index). Some have warned that prioritizing adults aged 65+ without working to remove barriers and promote equity could worsen existing racial disparities.^17^ Reitsma et al.^6^ concluded that equity-focused public policy is required to address disparities that have arisen during the pandemic. These studies highlighted the need for equity-focused modeling that explicitly captures the racial and ethnic demographics of the population, multigenerational households, demographic-informed workplace activity throughout the pandemic, and vaccine uptake as a function of racial/ethnic and age characteristics of the population.

We developed a COVID-19 transmission and disease progression model for North Carolina to compare vaccination prioritization strategies by total infections, severe outcomes, and disparities. In this analysis, we focused primarily on the first six months of the rollout in the United States, when vaccine availability was limited. The model incorporated risk of exposure based on mobility and disease severity among those at greater risk due to age or comorbidities. We evaluated prioritization strategies based on age (e.g., 65+), type of employment (e.g., non-medical essential workers), health risks (e.g., people with chronic conditions such as diabetes), and social vulnerability (e.g., historically marginalized populations). We also explored operational strategies such as susceptible-only distribution and increased uptake, which could be achieved by reducing barriers to access, building trust, increasing communication, or other methods^15, 17, 18^.

## METHODS

### Model Structure

We developed an agent-based extended SEIR simulation model with an embedded network structure where agents interact in households, peer groups such as workplaces or schools, and the community (Figure A1 and A2-supplement). We modeled the population of North Carolina using a ∼1:10 proportional representation of 1,017,720 agents. Census tract-level data^19^ was used to assign the distribution of individuals by race, ethnicity, age, and households of different sizes. Each agent is in one of (i) five age groups: children (0-4, 5-9, 10-18), adults (19-64), and older adults (65+), and (ii) four race/ethnicity groups: White (Non-Hispanic White), Black (Non-Hispanic Black), Other (Non-Hispanic Asian, American Indian or Alaska Native, Native Hawaiian or Other Pacific Islander, Other race, or multi-racial), and Hispanic (White Hispanic, Black Hispanic, Other Hispanic). Historically marginalized populations (HMPs) included Black, Hispanic, and Other. We incorporated SafeGraph mobility data at the census tract level, where we assumed essential workers were those working full-time away from home in January 2021^20^. Essential worker assignment followed the national racial/ethnic distribution^21^, which had a higher rate for non-whites (Figure A4-supplement). We included diabetes as a high-risk condition, which corresponds to increased risk of hospitalization^22^; with agent assignment based on the state-level prevalence specific to agents’ age and racial/ethnic group^23^. We modeled non-pharmaceutical interventions (NPIs) including face mask use and limited mobility over time (Figure 1). Using the model, we projected the disease spread for approximately 18 months, accounting for the Alpha variant strain’s spread in North Carolina. We compared various equity-driven vaccination prioritization strategies using outcome metrics of cumulative infections, hospitalizations, and deaths, and we assessed disparities using the age-adjusted differences of each outcome (with respect to the population of North Carolina) for each historically marginalized population relative to the White population.

**Figure 1.**
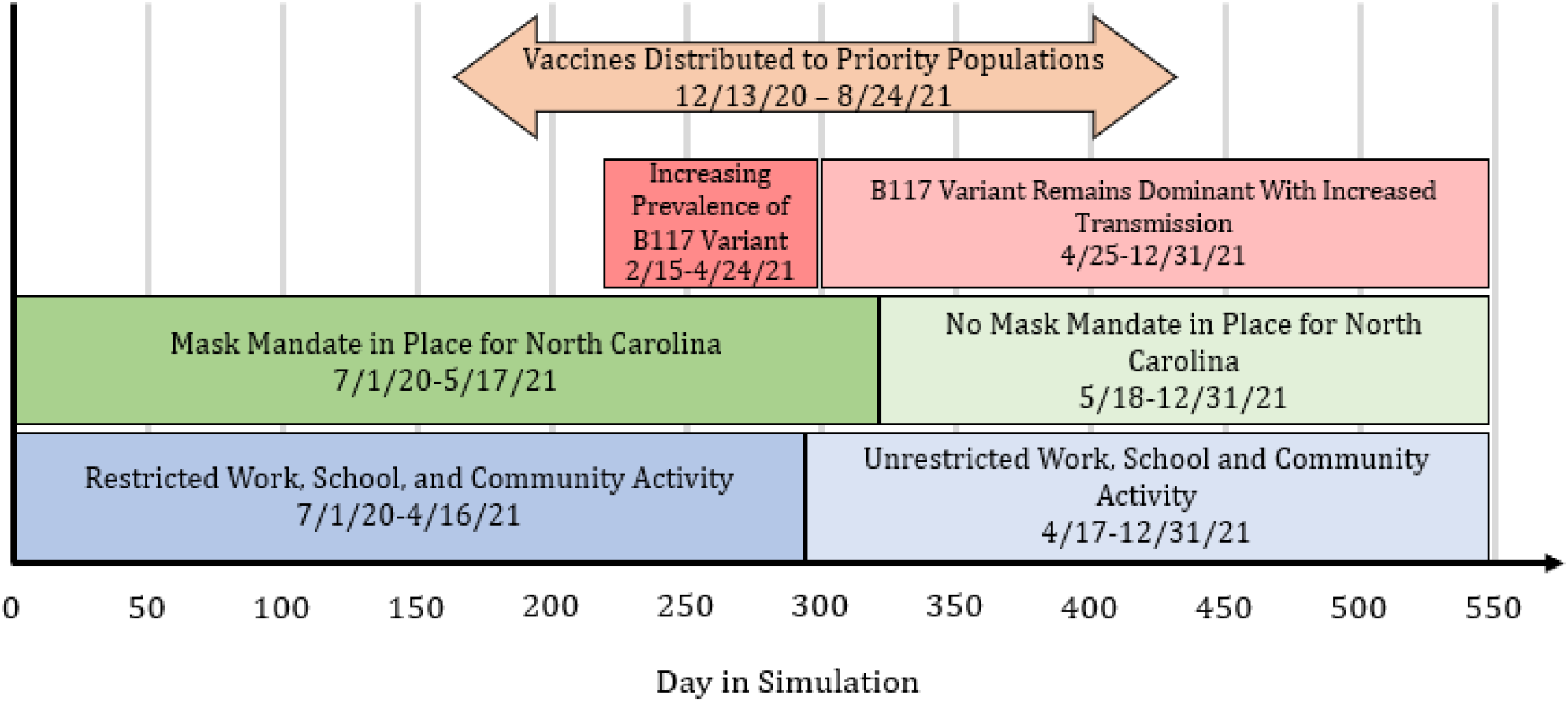
Caption: Simulation Timeline for NPI, Variant, and Vaccine Behavior Source: Agent-based simulation model timeline regarding NPIs, mobility, variants, and vaccination.

### Vaccination Prioritization and Distribution Scenarios

We modeled 29 vaccination-prioritization and distribution scenarios (Figure A13/A14/A15-supplement) considering a combination of (i) target vaccination groups, (ii) vaccine uptake, and (iii) susceptible-only distribution. Eight million doses were distributed at a uniform daily rate from January 10 until August 24, 2021. Vaccinations were administered to the 75+ and frontline populations beginning on December 13, 2020. In each scenario, the first dose distribution targeted a specific group until either 60% of the prioritized group had been vaccinated or uptake had been fully satisfied whichever occurred first, before extending eligibility to the next group.^24^ We assumed agents were eligible for a second dose 28 days after the first dose, with 15% attrition^25^. Vaccine efficacy occurred two weeks after a dose (70% after first, 90% after second). Vaccination implementation details can be found in the supplement.

#### Priority Population Combinations and Ordering Scenarios

The groups studied were people age 65+ (Age), adults with a high-risk condition, essential workers (Essential), HMPs, those that live in a low-income census tract, and the adult population for which there is no prioritization. Ordering scenarios consisted of two groups. After the second group was vaccinated, eligibility opened to all remaining adults. Here we focused on the Age and Essential priority groups (Age-Essential and Essential-Age). The baseline scenario for this study was the Age-Essential.^26^

#### Vaccine Uptake

The *baseline* vaccine uptake parameter captured a combination of an agent’s hesitancy to vaccinate and access to the limited supply (e.g., location/transportation accessibility, language barriers, etc.). Baseline vaccine uptake followed historical uptake trends from seasonal flu vaccine coverage.^27^ For age 65+, the uptake was 71.9%, 62.3%, 59.3%, and 71.9%, for White, Black, Hispanic, and Others, respectively. The corresponding uptake for ages 20 to 64 was 45%, 36.7%, 36.1%, and 45%, respectively.

Additional scenarios included increasing uptake to 100% for (i) HMPs and (ii) everyone. While 100% uptake may be unlikely in practice, this extreme scenario provided insightful results by establishing an upper bound for comparison. Since we assumed limited supply, most populations would not be fully vaccinated even under scenarios with 100% uptake.

#### Susceptible-only Distribution

Under the susceptible-only distribution strategy, doses are only given to susceptible agents. Operationally, this is comparable to administering an antibody test immediately prior to vaccination and only vaccinating individuals without antibodies. This maximizes the utility of the vaccine by leveraging the natural immunity of those previously infected.^14^

### Validation

The model has been used previously to answer questions regarding the impact of masks, school closures, testing strategies, and lifting of NPIs before full vaccination.^28-34^ We validated our model on cumulative lab-reported infections, hospitalizations, and deaths through April 15th, 2021, for the total population and subpopulations stratified by race/ethnicity or age, using estimates of lab-reported infections as described in the supplement. (Figure A9-supplement)

## RESULTS

### Baseline

At the time when vaccine distribution was assumed to begin, the simulated cumulative percentage of the population infected was approximately 18.6% with Black, Hispanic, Other, and White having 21%, 20.2%, 20.8%, and 17.6% age-adjusted infection rates, respectively. In the baseline vaccination prioritization scenario, we found the following simulated outcomes by December 31, 2021 at the state level: cumulative infections of 42.9%, 796 hospitalizations per 100,000, and 174 deaths per 100,000. This corresponded to the following outcomes for Black, Hispanic, Other, and White: 45.8%, 46.3%, 44.6%, and 42.4% age-adjusted infection rate; 838, 779, 748, and 691 age-adjusted hospitalizations per 100,000; and 181, 158, 152, and 135 age-adjusted hospitalizations per 100,000, respectively.

#### Effect of Prioritization and Ordering

At the state level, as shown in Figure 2a, the Age and Essential prioritizations led to a significant reduction in deaths (2.5% or 460 deaths) compared to no prioritization. Prioritizing Essential-Age significantly reduced infections (4.2%) compared to prioritizing Age-Essential, without significantly impacting the death rate. At the subpopulation level, Age-Essential prioritization reduced the White population’s death rate by 2.6% compared to no prioritization (Figure 2b vs 2d), but did not significantly reduce morbidity or mortality for any HMPs. Under Age-Essential prioritization (Figure 2c), greater morbidity and mortality continued for HMPs compared to the White population, 8.1%, 9.2%, and 5.3% more infections and 34.4%, 17.1%, and 13.2% more deaths for Black, Hispanic, and Other populations, respectively.

**Figure 2.**
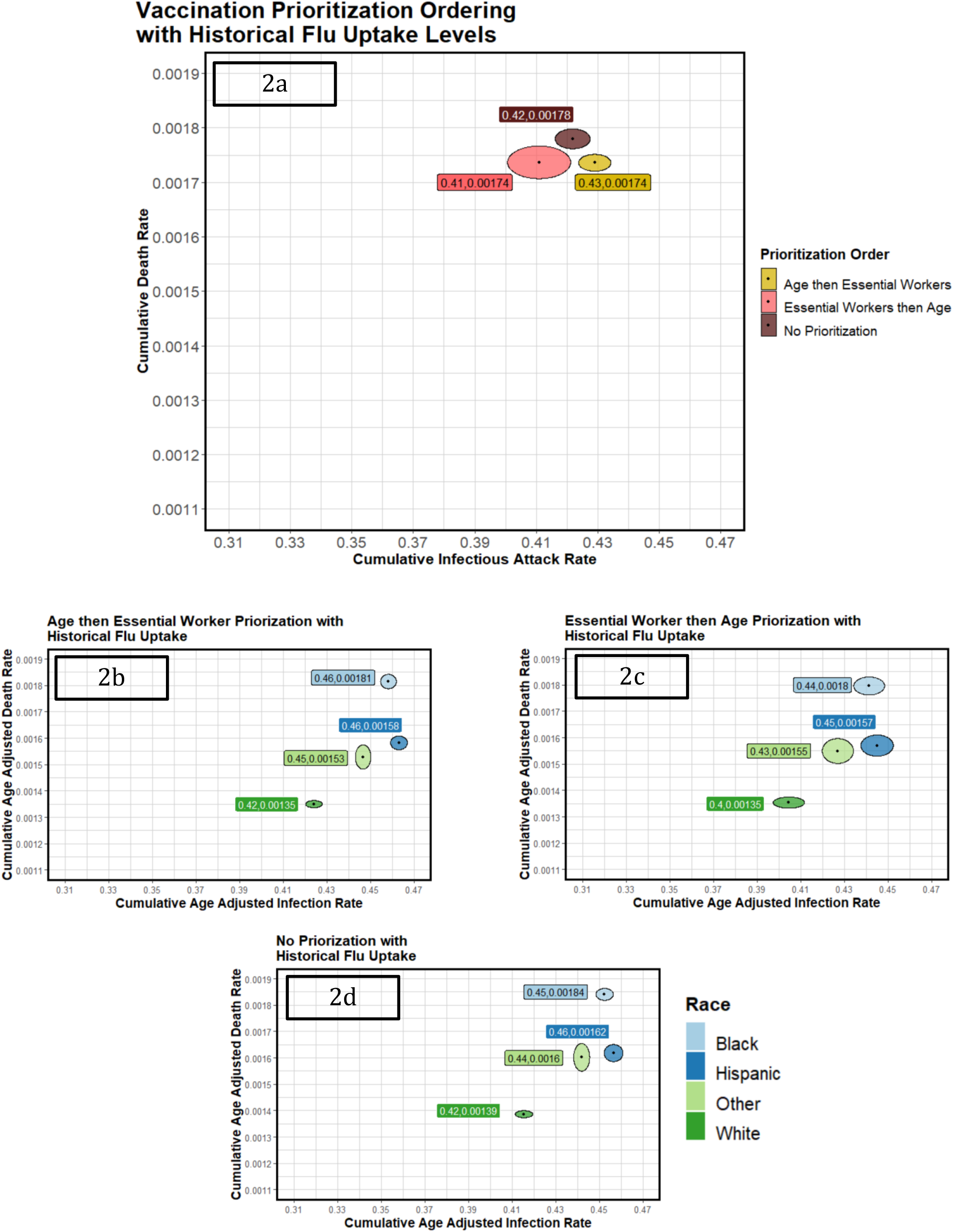
Caption: Impact of Prioritization Under Historical Flu Vaccine Uptake at the State and Subpopulation Levels. Source: Data shown is generated from the agent-based simulation model across 45 replications. NOTES Each panel shows the average age-adjusted infection attack rate (x-axis) and the average age-adjusted death rate (y-axis) with 95% confidence intervals represented with ovals, with scales consistent across all similar figures. Panel (a) provides state level results, and panels (b)-(d) provide results stratified by race/ethnicity.

Six additional prioritization scenarios are shown in Figure A13 in the supplement. Among the scenarios, Essential-Age reduced morbidity the most for the state level population and at the subpopulation level. Age-Essential and Essential-Age reduced mortality the most for the state level population and at the subpopulation level with historical flu vaccine uptake.

#### Effect of Vaccine Uptake and Susceptible-Only Vaccination on Outcomes

At the state level (Figure 3a), increased uptake in the HMPs reduced deaths by 2.6% or 472 deaths prevented and did not significantly increase the infection rate compared to baseline uptake. At subpopulation level, we saw that increased uptake corresponded to significant reductions in both morbidity and mortality for the Black, Hispanic and Other populations: 9.4%, 8.9%, and 6.7% reduction infections and 18.9%, 17.8%, and 12.5% reduction in deaths, respectively (Figure 3c). Compared to the White population, the Black, Hispanic and Other populations had 9.3%, 7.9%, and 9.0% fewer infections. Despite the reduced infection rate, the Black population had 2% more deaths while the Hispanic and Other populations had 9.8% and 7.2% fewer deaths under the increased uptake scenario compared to the White population. Figure 3a shows that susceptible-only distribution could lead to additional reduction in morbidity and mortality, at the state level, and for each subpopulation. There was a 4.6% and 3.8% reduction in infections and deaths, respectively, relative to the historical flu vaccine uptake scenario. At the subpopulation level, Figure 3d shows that susceptible-only distribution led to a significant reduction in morbidity for the Black, Hispanic, Other and White populations: 3.5%, 4.2%, 3.9%, and 5.1%, respectively, and a significant reduction in mortality for the Black and White populations 4.0% and 4.2%, respectively. For Age-Essential, there was no reduction of disparities associated with susceptible-only distribution alone.

**Figure 3.**
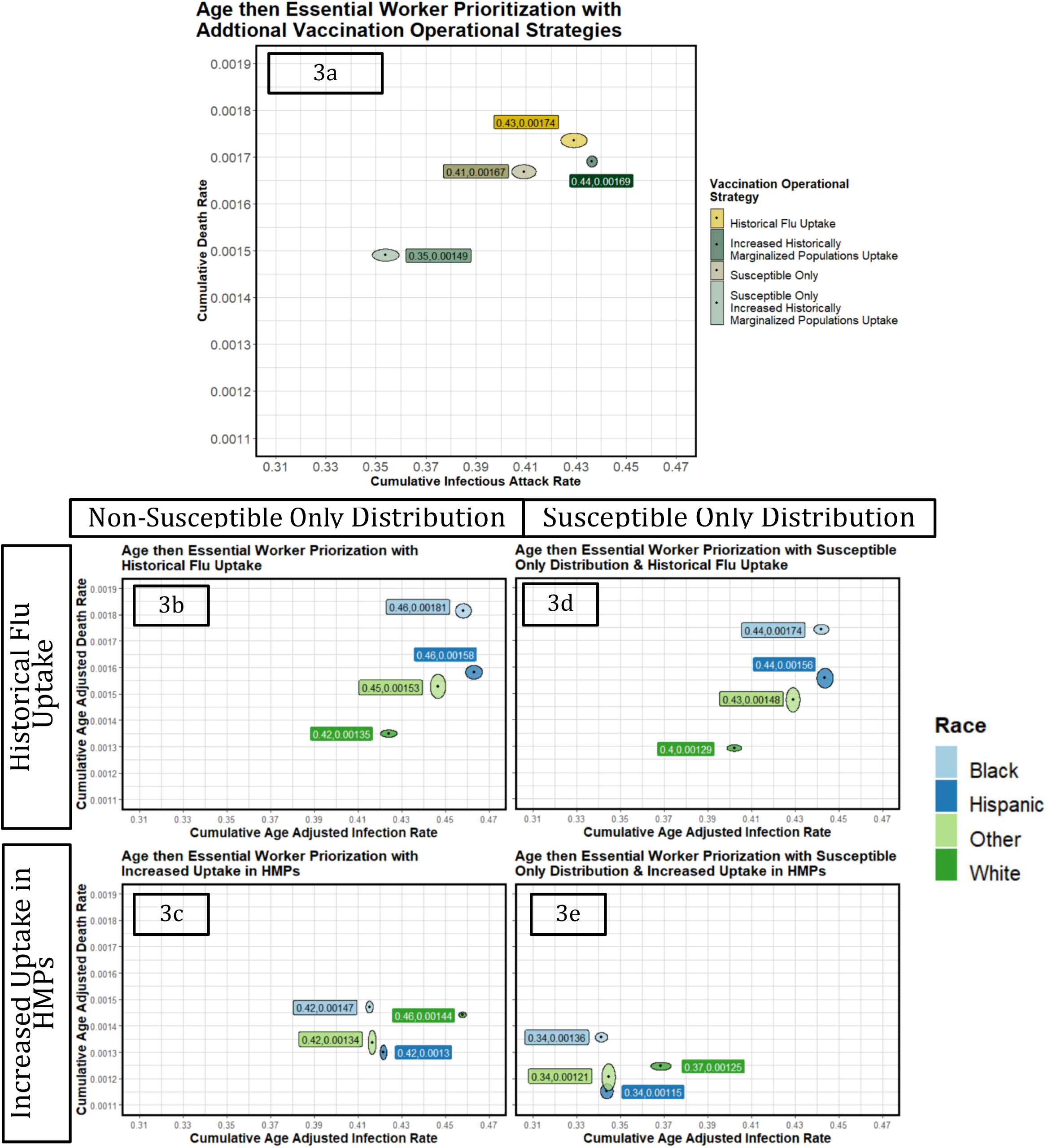
Caption: Impact of Vaccine Uptake and Susceptible-Only Distribution at the State and Subpopulation Levels Source: Data shown is generated from the agent-based simulation model across 45 replications. NOTES Each panel shows the average age-adjusted infection attack rate (x-axis) and the average age-adjusted death rate (y-axis) with 95% confidence intervals represented with ovals, with scales consistent across all similar figures. Panel (a) provides state level results, and panels (b)-(d) provide results stratified by race/ethnicity.

When susceptible-only distribution and increased uptake in HMPs were operationalized simultaneously, the greatest reduction of morbidity and mortality relative to the baseline scenario was observed. At the state level, there was a 13.8% and 14.1% percent reduction in infections and deaths, respectively (Figure 3a). By subpopulation, the Black, Hispanic, Other and White populations’ morbidity and mortality were reduced by 25.4%, 25.7%, 22.8%, and 13.1%, and 25.2%, 27.2%, 21.1%, and 7.6%, respectively (Figure 3e). Despite the large reductions, the Black population continued to have the highest death rate.

Results for 16 other scenarios are in Figure A14/A15 of the supplement.

Figure 4 shows the equity gap pre-and post-vaccine administration. Under vaccination prioritization scenarios without increased uptake in HMPs, the equity gap increased significantly relative to the pre-vaccine values. When uptake is increased to 100% in HMPs, the Hispanic and Other populations achieved lower death rates than the White population. However, the Black population still faced significant disparities in the age-adjusted death rate relative to the White population under all scenarios. Figure A11 and A12 in the supplement show the corresponding graphs for cumulative age-adjusted infection and hospitalization rates, respectively.

**Figure 4.**
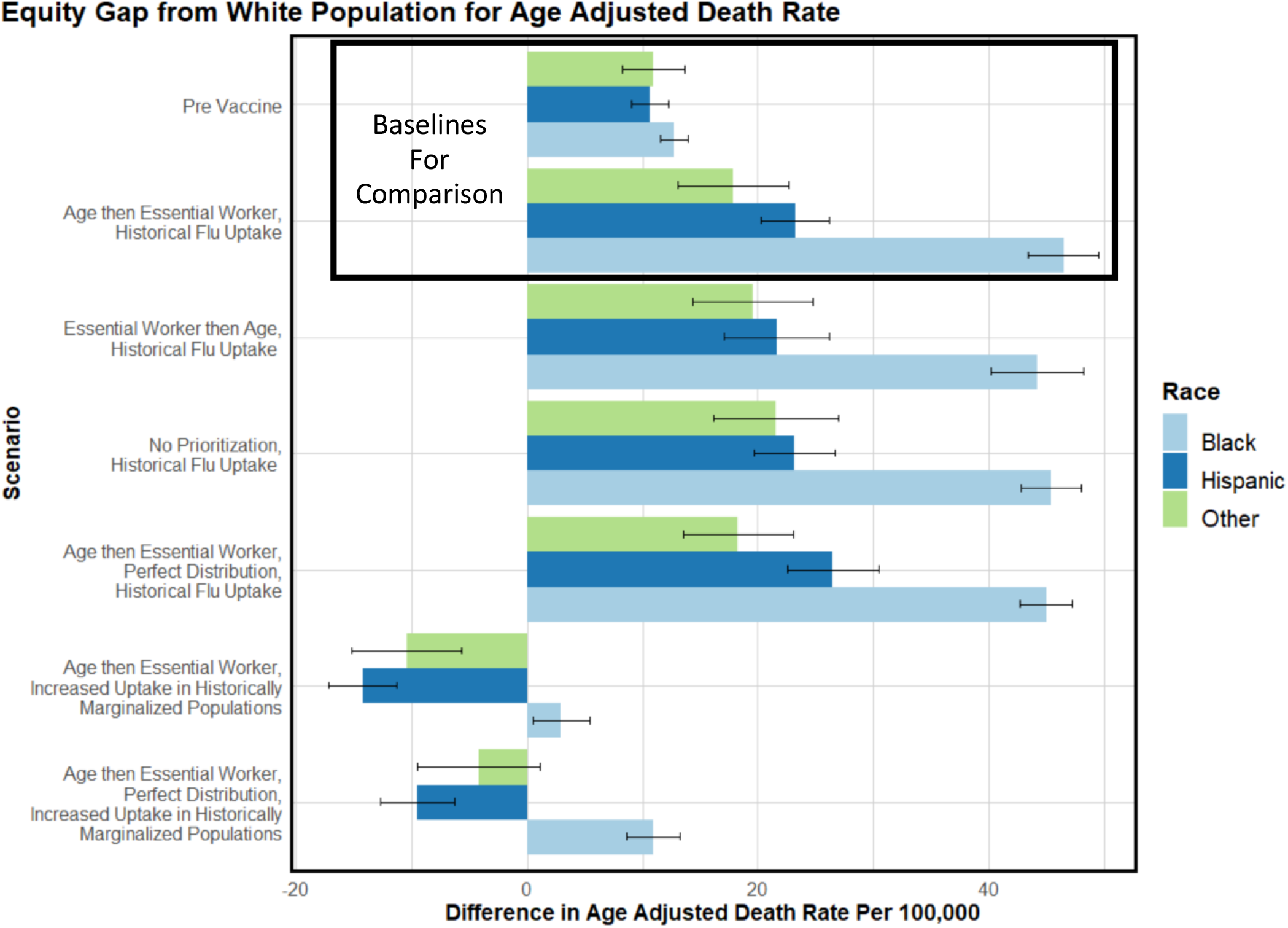
Caption: Equity Gap from the White Population for Each Historically Marginalized Population by Vaccination Scenario Source: Data shown is generated from the agent-based simulation model across 45 replications. NOTES Equity gap is defined as the difference in cumulative age-adjusted death rates between the White and each historically marginalized population. The y-axis indicates the vaccination scenario. Figure contains the corresponding 95% confidence intervals.

## DISCUSSION

Using a simulation-based model, we compared several COVID-19 vaccination strategies. We found that prioritization schemes that did not incorporate increased uptake in historically marginalized populations did not reduce disparities and may further exacerbate pre-vaccine disparities. For infections, hospitalizations, and deaths, the results showed an increased age-adjusted equity gap for historically marginalized populations, unless there is increased uptake. At the extreme, we found that, even with universal uptake, the Black population continued to have a higher post-vaccine death rate than the White population. Although equity is one of the tenets of vaccine distribution,^4, 24, 26^ the current distribution strategy outlined by federal agencies is insufficient to reduce disparities between racial and ethnic groups that existed pre-vaccination.

Disparities in COVID-19 outcomes may persist and even worsen in historically marginalized populations due to systemic biases that increase the risk of infection and severe disease and lower historical uptake,^1-3, 5, 6, 35^ some of which were captured in our model. Historically marginalized populations face greater workplace exposure due to their disproportionate essential worker status in the model (Figure A4-supplement)^1^. The Hispanic population has larger average household sizes (Figure A5-supplement),^19^ and the Hispanic and Black populations have a younger average population (Figure A5-supplement).^19^ These factors correspond to more interactions in the model. The Black population has a higher rate of diabetes for both 20-64 and 65+ adults (Figure A6-supplement).^23^ The diabetes rate in combination with essential-worker mobility, contributes to the high death rates relative to other populations and the equity gap observed pre-vaccination (Figure 4). When risk factors are coupled with lower historical uptake^27^ and biases that result from age prioritization in a population where Whites comprise the majority of the older population,^19^ historically marginalized populations may face increasing disparities (Figure 4). In addition to these risk factors, others such as systemic racism likely exacerbate disparities in morbidity and mortality.

Our findings are consistent with previous studies that show prioritizing the older adult population is important for reducing mortality at the state level ^6, 9-11^. Further, our study suggests increasing uptake in historically marginalized populations is critical for reducing disparities in COVID-19 morbidity and mortality. Under constrained supply, our model shows that leveraging natural immunity through susceptible-only distribution is critical to have in combination with increased uptake to reduce morbidity and mortality for all subpopulations. These operational vaccination strategies would not have required additional vaccine supply, yet they could have averted 830K infections and 2,700 deaths compared to the baseline. Such strategies, however, might require additional laboratory infrastructure to confirm existing immunity, which may not be feasible, especially in rural and resource-limited areas. To improve uptake, both access and vaccine hesitancy must be addressed with equity-focused public health policy.^6,15,36,37^ This could take the form of mobile vaccine clinics, removing registration barriers, providing multilingual communication on registration and vaccine safety, extended operational hours, paid time off, or travel expense compensation^4, 37-41^. Additional approaches could include interpersonal communication with healthcare professionals that focus on personal benefits of receiving a vaccine and working to dispel misinformation regarding the vaccine’s safety and efficacy ^36,42,43^.

Significant equity gaps existed for historically marginalized populations prior to vaccine rollout. Equity-focused public health policy needs to extend beyond the scope of the pandemic to address the root causes of these disparities. These include ensuring equitable access to healthcare resources and taking action to reduce the prevalence of high-risk conditions such as diabetes. Establishing more equitable public health policies now will better protect vulnerable populations in the future as uncertainty remains due to the emergence of new and potentially more infectious variants of SARS-CoV-2 ^44-46^. Equitable policy is particularly important when planning for potential booster shots, where access barriers will increase morbidity and mortality within historically marginalized communities of color and communities with low uptake.

### Limitations

Validation shows the model underestimated disease burden within the Hispanic community. This is due in part to data limitations surrounding the migrant worker population. As a low estimate, 150,000 migrant farm workers come to North Carolina each growing season, with 94% being native Spanish speakers ^47^. Nationally, 53% of migrant workers are undocumented which leads to underreporting in census data^47^ and leads to a misrepresentation of the population within the simulation. The age bracket definitions are also a limitation within the model. For the adult population, we are not able to capture the workplace mobility or community interaction differences. This also limits the assignment of diabetes within the population. The older adult population does not have a workplace peer group, which limits our ability to capture disease spread and the racial/ethnic and comorbidity-based disparities that arise in this population. The variant is modeled by increasing the transmissibility of the disease, rather than introducing a competing strain into the population. This implementation may overestimate the impact of the variant on disease spread. Additionally, there is limited understanding of the variant’s prevalence in the population, as genomic surveillance in North Carolina is limited ^48^. Finally, we assume masking ends on a particular date whereas in reality, people may continue to wear masks voluntarily and through workplace or school mandates. As a result, our model may overestimate infections (Figure A9-supplement).

## CONCLUSIONS

Our analysis suggests that the racial and ethnic disparities in COVID-19 morbidity and mortality that existed before the availability of effective vaccines could be exacerbated by vaccination prioritization strategies that do not directly increase uptake within historically marginalized populations. Across all scenarios, we found that prioritizing older adults had the greatest impact on reducing mortality, while prioritizing essential workers had the greatest impact on reducing morbidity. Disparities in disease burden could only be reduced through targeted strategies to increase uptake. It is critical to consider public health policies that emphasize equity in planning for vaccine boosters and mass vaccination strategies for future pandemics.

## Data Availability

This manuscript uses publicly available data from the US Census, published estimates of disease parameters, data obtained from SafeGraph on mobility across communities, and estimates obtained from the Covid-19 Case Surveillance Restricted Access Detailed Data.

## Author Contributions

Drs. Ivy, Mayorga and Swann had full access to all the data in the study and take responsibility for the integrity of the data and the accuracy of the data analysis.

*Concept and design:* Ivy, Mayorga, Rosenstrom, Swann.

*Acquisition, analysis, or interpretation of data:* Ivy, Mayorga, Rosenstrom, Mele, Patel, Boyce, Smith, Johnson, Delamater, Hasmiller Lich, Swann.

*Drafting of the manuscript:* Mele, Rosenstrom, Ivy, Mayorga, Boyce, Swann.

*Critical revision of the manuscript for important intellectual content:* Ivy, Mayorga, Patel, Rosenstrom, Mele, Keskinocak, Boyce, Hassmiller Lich, Smith, Delamater, Swann.

*Statistical analysis:* Mele, Rosenstrom, Ivy, Mayorga, Swann.

*Obtained funding:* Patel, Keskinocak, Ivy, Mayorga, Hassmiller Lich, Swann.

*Administrative, technical, or material support:* Patel, Ivy, Keskinocak, Mayorga, Boyce, Smith, Johnson, Swann.

*Supervision:* Patel, Ivy, Mayorga, Keskinocak, Hassmiller Lich, Swann.

## Disclaimer

The content is solely the responsibility of the authors and does not necessarily represent the official views of the NIH, CSTE, CDC, or the universities employing the researchers.

