## Supplemental Material for "Can Vaccine Prioritization Reduce Disparities in Covid-19 Burden for Historically Marginalized Populations?"

SUPPLEMENT

1. **SEIR Compartment Model**

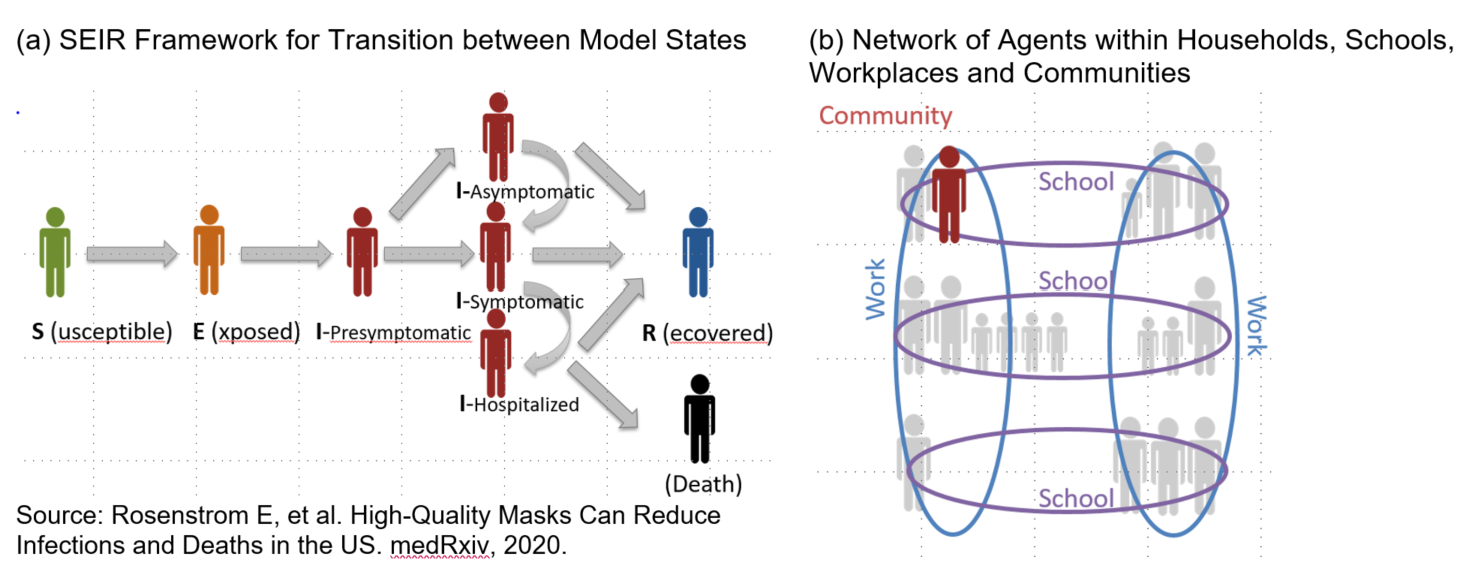

**Figure A1**: Extended SEIR Compartment Model

Source: Rosenstrom E, et al. High-Quality Masks Can Reduce Infections and Deaths in the US. *medRxiv*, 2020. **doi:** https://doi.org/10.1101/2020.09.27.20199737^1^.

Disease progression is modeled as an extension of a Susceptible-Exposed- Infectious-Recovered (SEIR) model (Figure A1). Each agent exists in one of the following states: susceptible (S), exposed (E), pre-symptomatic (IP), asymptomatic (IA), symptomatic (IS), hospitalized (IH), dead (D), or recovered (R). Each individual begins in the susceptible disease state unless they were assigned one of the exposed, dead, or recovered states during initialization (seed). When an agent enters the recovered or dead states they do not leave that state. Once a susceptible individual who is in close contact with an infected individual is identified, their disease progression begins, and they enter the Exposed state. An agent’s likelihood of traversing one path as opposed to another (i.e., the transition from IS to IH or from IS to R) is dependent on transition probabilities that may also differ by age and health status. The transition from IS to IH is also dependent on if the agent has a high risk condition. Only agents from the symptomatic infection state can be hospitalized, which represents a severe case, and only severe cases can lead to death. The length of time spent in any disease state is a random variable, with the time in all intermediate disease states being governed by defined probability distributions. The list of input parameters and associated references is provided in Figure A7. Additional details are available in the supplement of [censored for double-blind review]^2^ .

The reproductive number R0, indicates the infectivity of the virus at the beginning of the pandemic and before any interventions are initiated. This value is directly related to the transmission rate (denoted as β). Other critical parameters that are relevant to disease transmission are the proportion of transmission that occurs outside the household, γ, and the proportion of transmission that occurs in the community δ. For disease-state specific transmission, θ, is the proportion of transmission that occurs in IP or IA health states. This leads to ω, which is the proportion of transmission by patients who are never symptomatic.

Compared to the values for influenza used in ^3^, the proportion of asymptomatic transmission is higher. In comparison to the model parameters used in previously validated work for North Carolina^4^, this model incorporates a time-varying transmission rate.

2. **Agent Network Structure**

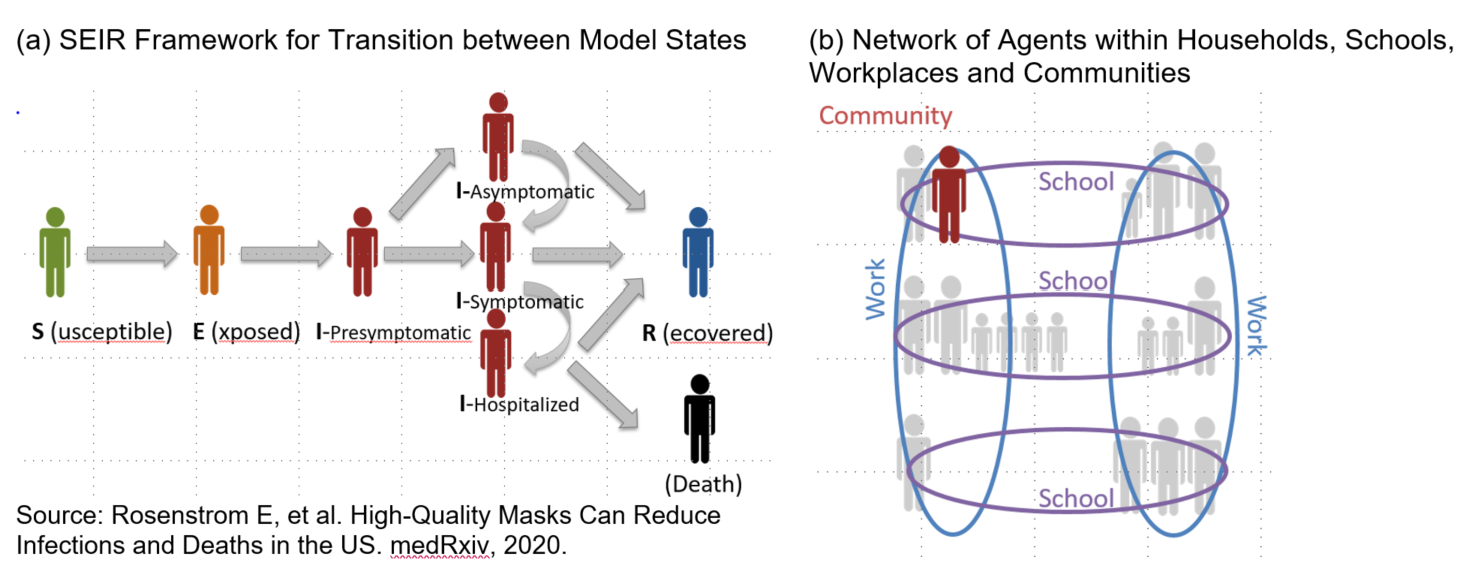

**FigureA2**: Agent Interaction Network

Source: Rosenstrom E, et al. High-Quality Masks Can Reduce Infections and Deaths in the US. *medRxiv*, 2020. **doi:** https://doi.org/10.1101/2020.09.27.20199737 ^1^.

A network exists in the model to simulate agents’ interactions in the household, peer group and community (Figure A2). The peer group represents a school for children and a workplace for adults. The implementation of each peer group is fundamentally the same. Communities are composed of all agents in the same census tract. The model represents North Carolina's statewide population of 10,490,000 people using 1,017,720 agents. The representative agent population were created using the US Census data, including household size (1 to 6), the presence of children^5, 6^, and the race/ethnicity of the household (with reporting for Black Only, White Only, Hispanic not White or Black, etc.) Individuals in the household are assigned the same race/ethnicity, independent of other factors. Using data tables at census tract level that were publicly available, individuals were assigned to age in five categories (age 0 to 4; age 5 to 9; age 10 to 19; age 20 to 64; age 65 and greater)^5^, with the head of the household always 18 or older. This limitation implies that our model is capturing less heterogeneity than reality. Multigenerational households exist within the model. As a surrogate for high-risk conditions, we determine the conditional probability of an agent having diabetes given their age and race/ethnicity using data regarding the statewide prevalence of diabetes^7^. During the day, all agents 5 to 19 interact in peer groups (“schools”), and agents 20 to 64 interact in peer groups (“work”) according to commuting patterns. Workflow data indicates the people who live in a census tract and the percentage who work in each of the other census tracts^8^, and is used to create workplace peer groups that contain agents from different census tracts. When schools are closed (or hybrid), children stay at home (or stay home every other day). All agents interact with household members at night.

For adult agents, age 20 to 64, workplace peer group activity is determined using SafeGraph^9^ mobility data to find the proportion of agents going into work during the day. SafeGraph is a data company that aggregates anonymized location data from numerous applications in order to provide insights about physical places, via the Placekey Community (Placekey.io)^9^. To enhance privacy, SafeGraph excludes census block group information if fewer than five devices visited an establishment in a month from a given census block group. SafeGraph tracks device location data and classifies each as working full-time if that device spent greater than 6 hours at a location other than their home geohash-7 during the period of 8 am - 6 pm in local time. The data indicate the count of devices classified as working full-time. Due to data sparseness, census tracts with the same Rural-Urban Commuting Area (RUCA)^10^ classification, which assess the urbanicity of the census tract and the same state-level income quartile are aggregated and assigned the same average mobility. Adult agents are assigned a workplace threshold number generated from a uniform distribution that is dependent on the agent's race/ethnicity at the beginning of the simulation. This number is compared to their census tracts' mobility every month to determine if the agent will have an active peer group. The upper bound of uniform distribution is race/ethnicity dependent to appropriately fit the national race/ethnicity distribution of essential workers seen in Figure A4. This workplace peer group activity implementation ensures that some agents are consistently working during the pandemic, while others are either leaving or returning to work corresponding to the mobility data. This decrease in workplace activity reduces the infectivity of the workplace. Figure A3 shows SafeGraph^9^ mobility for North Carolina.

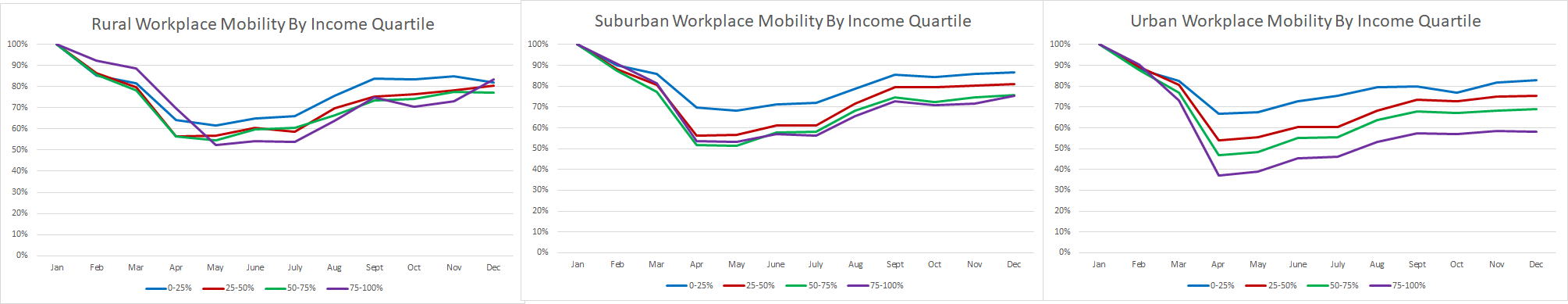

**Figure A3**: SafeGraph^9^ Workplace Mobility for 2020, summarizing workplace mobility by income quartile for urban, suburban, and rural.

| Race/Ethnicity | Proportion of Population^11^ | Proportion of Essential Workers ^11^ |
| --- | --- | --- |
| Black | 21.8% | 15% |
| Hispanic | 9% | 21% |
| Other | 4.2% | 9% |
| White | 65% | 55% |

**Figure A4**: Agent Population By Race/Ethnicity and Distribution of Essential Workers

Safegraph^9^ and Google mobility^12^ were reviewed for non-workplace related mobility. To model this mobility, we assume community mobility in the simulation model follows the same pattern as workplace mobility, but where the deviations from the pre-pandemic mobility are scaled down. The mobility was scaled by a factor of 2. For example, if the mobility is at 80% then the community factor is 90% of the original community value (specific calculation for new Community parameter: (1-((1-0.8)/2) * (community_parameter = 0.23) = 0.207). The scaling factor of 2 was experimentally determined. This implementation models a reduction in interactions and agents in the community as a reduction in the community infectious hazard.

3. **Demographics**

The census data used to construct the simulation network uses independent data sources e.g., data only constructs households based on the census tract, not the agent’s race and the census tract. Since the data sources that inform the attributes of agents are independent, when examining their intersection with race/ethnicity there is less heterogeneity than would be seen in reality. However, we are able to capture the correct directionality of the trends on aggregate due to the granularity of the data. Various demographic properties are shown in FigureA5 and FigureA6. The average age is calculated using the median age value for each bucket multiplied by the proportion of the population in that age bucket. We take 75 as the value for 65+ agents.

At the state-level the resulting average age is highest in the White population followed by the Black and Hispanic populations. The resulting average household size is largest for the Hispanic population followed by the Black and White populations. These results are consistent with the directionality of national trends^13, 14^.

| **Simulation Statistic** | **Black** | **Hispanic** | **White** |
| --- | --- | --- | --- |
| Average Age | 37.96 | 37.9 | 39.31 |
| Average Household Size | 2.37 | 2.43 | 2.36 |

**Figure A5**: Agent Population Age and Household size by Race/Ethnicity

Figure A6 shows the distribution of diabetes within the population. Diabetes is a key high-risk condition which is assigned to agents in the population based on race and age using state level prevalence data^7^. The older adult Black population has the highest proportion of agents with diabetes. The large age-bucket representing adults (20-64) limits the distribution of diabetes. There are differences in the distribution in diabetes within this age-bucket.

| **Race/Ethnicity** | **Age** | **Likelihood of Diabetes In Simulation** |
| --- | --- | --- |
| Black | 20-64 | 0.082 |
| Black | 65+ | 0.339 |
| Hispanic | 20-64 | 0.066 |
| Hispanic | 65+ | 0.202 |
| Other | 20-64 | 0.084 |
| Other | 65+ | 0.229 |
| White | 20-64 | 0.084 |
| White | 65+ | 0.229 |

**Figure A6**: Agent Population Likelihood of Diabetes by Age and Race/Ethnicity

4. **Extended Methodology on Vaccination and Operational Policies**

During the phase of limited resources, vaccines are allocated to successive priority populations, e.g., essential workers, people 65+, historically marginalized populations, or people with high-risk medical conditions, and the general population. The vaccine is also shared among those receiving their first dose and those receiving their second dose. An uptake parameter is defined to determine how many people in a given population will take the vaccine when it is available; the parameter incorporates demand and access. We also allow for the possibility that vaccines may be distributed to the population that is still susceptible (i.e., has not had the disease already). The modeling also incorporates adding the next priority population. Details are provided below on each of these elements.

**Eligible Priority Population:** The population eligible for first-dose at a specific time includes any persons with a positive uptake value, who are in previous or currently eligible populations, and who have not had their first dose. The population eligible for a second-dose at a specific time includes all people who have had a first-dose at least 28 days previously and have not yet had their second dose.

**First-Dose vs Second-Dose Vaccination:** For a given vaccine dose, it is equally likely to be given to the first or second-dose population. Within the selected dose population, it is equally likely to be given to any agent. If no agents are in the first-dose population, all vaccines are given to the second dose population, and vice versa.

**Priority Group Switching Criteria:** In each scenario, the distribution of the first dose targets a specific group, with a shift to the next eligible priority group when either at least 60 percent^15^ of the current full population priority group has been vaccinated or the entire eligible priority group has been vaccinated, whichever occurs first. If there are agents in the current eligible priority group at the time the switching criteria is met, they are added to the next eligible priority population group. Once all of the agents in the general vaccination population have been vaccinated, no more agents will receive a first dose.

**Susceptible-Only Vaccination:** Under the susceptible-only operational strategy, doses are only given to susceptible agents. The susceptibility of the agent is known with certainty as the disease state of each agent is tracked throughout the simulation. There would be uncertainty in the outcome of the test (false negative/positive). This maximizes the utility of the vaccine by leveraging the natural immunity of those previously infected.^16^

**Essential Worker Prioritization Group:** The essential worker prioritization group is chosen immediately prior to vaccination distribution beginning in the simulation. The group is chosen from the subset of the population that are adults and have a currently active workplace peer group. We select a portion of the working population to account for frontline workers who were the first to receive the vaccine. These agents are guaranteed to have an active workplace peer group for the entire simulation.

**Non-Pharmaceutical Interventions (NPIs) Assumptions**

We use data to represent behaviors associated with non-pharmaceutical interventions such as those directed by state or local policies. As discussed earlier mobility data is used to model reduced workplace peer group activity, and schools have hybrid attendance. We assume that masks reduce infectivity and susceptibility by 50% when they are worn^4^. Masks are originally distributed throughout the population at the beginning of the simulation following Facebook mask compliance data^17^. The distribution is dependent on the urbanicity of the census tract as defined by its RUCA classification. If an agent is assigned a mask it is worn only in the peer group and community settings. If an agent is not assigned a mask, they will never wear a mask.

We make assumptions about the behavior of agents to quantify the impact of the statewide policy decisions that occurred in April and May in North Carolina. For agent mobility, on April 17, 2021, all children and working age adults return fully to their peer groups, and all agents return to the community, coinciding with the approximate time when statewide mobility increased and students returned to school in North Carolina^18, 19^. On May 18, 2021, all agents remove their masks for the remainder of the simulation. This corresponds to the governor of North Carolina’s policy that extended the CDC guidance lifting masking requirements for vaccinated individuals to lifting mask requirements for everyone^20^.

**Modeling Variants**

Many variant strains of COVID-19 have appeared in the United States during the first half of 2021. We model the variant strain B.1.1.7 (Alpha) spread into North Carolina which began in early February and reached a dominant status in late April accounting for 58% of the total infections according to the CDC^21^. A recent study in the United Kingdom indicated that this strain is 1.3-1.7 times more transmissible than the original strain of COVID-19^22^. Per our model validation procedure, we remain on the low end of this range and mode

l a variant strain with 1.3 times increased transmission. The variant is implemented in the model by increasing the transmission rate linearly over 10 weeks. The final transmission rate is the weighted average of the original transmission rate and 1.3x the original transmission rate with weights corresponding to the proportion of cases found in testing surveillance. This weighted average transmission rate remains for the remainder of the simulation.

**Outcomes Information**

For each scenario, we compute the following outcomes: infectious attack rate, hospitalization rate, and death rate. To compare fairly across populations, we also calculate average age-adjusted outcomes by race/ethnicity where we use the total population of North Carolina as the reference population. Disparities are quantified by the difference in age-adjusted rates (cases, hospitalizations and deaths) between the White population and each historically marginalized population.

We ran 45 replications for each scenario, quantifying the variability (standard deviation or standard of error with a two-sided 95% confidence interval, 𝛂 = 0.05) for each outcome measure associated with that scenario. This allows us to determine whether a particular prioritization or operational scenario has a different outcome than a comparative scenario.

**Estimation of Cases: Seeding/Lab Multiplier**

Our goal was to seed the model for a point in time with cases that were infectious or hospitalized, with a distribution of these cases across each census tract.

It is well-known that the number of true cases is much higher than the number of lab-reported cases, and that it changes over time^23, 24^. From early to mid-2020 it was estimated that approximately 1 in 7.1 infections were reported nationally whereas recent estimates through early 2021 suggest 1 in every 4.3 infections were reported^23, 24^. It likely also differs by state, e.g., New York may have more under-reporting than NC since it experienced its first large surge when testing was limited. The reported values of deaths and hospitalizations are much closer to true values than reported infections. Recent estimates suggest that approximately 1 in every 1.8 COVID-19 related hospitalizations were reported ^24^and the total reported COVID-19 death count falls within the range of deaths approximated from excess mortality data^25^.

Estimating True Infections:

To estimate the number of cumulative infections in the state by a given date, we divide the cumulative deaths 21 days after the specified date with an overall true infection fatality rate of 0.5%^4, 26, 27^. This gave us the expected number of infections that would have been needed to result in the reported number of deaths. 21 days represents a three-week time lag corresponding to the length of time between exposure to death^28^. From this value of the expected number of infections, we derived a time-dependent lab multiplier to use for seeding and validation.

We find that our lab multiplier has the highest values in March and April and decreased to an average of 2.8-3 towards the end of our validation period, which is consistent with the findings of the CDC^23, 24^ that show national values decreasing by early 2021.

Active cases, hospitalizations, deaths, and recovered individuals are seeded from July 1, 2020, which corresponds to the beginning of the simulation.

Deaths: Deaths are seeded following the age distribution of reported deaths through July 1, 2020.

Hospitalizations: The North Carolina Department of Health and Human Services (NC DHHS) ^29^ reports daily admitted and current COVID-19 hospitalizations by hospital region, where each region serves multiple counties. To allocate region-level hospitalizations to each county, we estimated county-level hospitalizations using a bi-level apportionment algorithm. We first estimate county-level daily admitted hospitalizations based on the number of daily cases in each county from four to six days prior and secondly, we estimate county-level currently hospitalized based on the number of estimated daily admitted hospitalizations from up to two weeks prior.

Recovered: Following initial CDC guidelines on quarantine recommendations^30^, we assumed that cases remained active, or infectious, for two weeks. We estimated the number of cumulative recovered cases by a given day by summing the cumulative number of estimated infections from two weeks prior, by applying our lab multiplier to the cumulative number of reported cases and subtracting the cumulative number of reported deaths.

Active cases: To estimate the number of active cases on a given day we estimated the number of true infections and subtracted the number of deaths, approximate recovered cases, and current hospitalizations. Hospitalized cases are considered isolated in the model and therefore, do not contribute to the number of infectious agents on a given day. We allocate all active cases to the exposed state upon the beginning of the simulation.

The simulation seeds for all deaths, hospitalizations, recovered, and active cases are distributed to each census tract using an apportionment algorithm based on population size and scaled to our generic population size. We used a similar approach to estimate and seed the model at different points in time, including March 24, 2020, and examined the results for values during the initial warm-up and the final values, compared to both NC DHHS^29^ and NY Times data^31^, as described in the validation section below.

5. **Model Parameters**

| **Parameter** | **Estimates** | **References** |
| --- | --- | --- |
| Exposed (E) Duration | Weibull with mean 4.6 days | [^32^, ^33^] |
| Pre-symptomatic (IP) Duration | 0.5 days | [^32^] |
| Hospitalized (IH) Duration | Exponential with mean 10.4 days | [^32^, ^34^] |
| Symptomatic (S) Duration | Exponential with mean 2.9 days | [^26^] |
| Symptomatic-Asymptomatic Duration Ratio | 1.5 | [^32^] |
| Probability of Symptomatic (from IP) | 0.50-0.82 | [^35^,^36^,^37^,^38^,^39^] |
| Probability of Hospitalization without Diabetes | 0.0033 for age 0-19;  0.018 for age 20-64;  0.11 for age 65+ | [^28^, ^40^, ^41^] |
| Probability of Hospitalization with Diabetes | 3 times rates above | [^42^] |
| Probability of Death (from IH) | 0 for age 0-19;  0.0515 for age 20-64;  0.3512 for age 65+ | [^28^, ^40^] |
| R_0_ | 2.4 | [^43^,^44^,^45^] |
| R_0_ Variant | 3.12 | [^22^] |
| β transmission rate | 1.12 | [^43^] |
| θ (probability IP to IA) | 0.48 | [^33^] |
| ω (proportion infections by IA) | 0.24 | [^33^] |
| γ (proportion of transmission that occur outside households) | 30% | [^3^, ^2^] |
| δ (proportion of infections outside households that occur in community) | 0.23 | [^2^]; Calibration |
| FlatImportRate | 45 | [^2^]; Calibration |

**Figure A7**: Key Model Parameters

**Vaccine Parameters**

| **Parameter** | **Estimates** | **References** |
| --- | --- | --- |
| Vaccine Efficacy Dose 1 | 0.7 | Johnson and Johnson [^46^] |
| Vaccine Efficacy Dose 2 | 0.9 | Pfizer [^46^] |
| Time Between Doses | 28 days | Pfizer/Moderna [^47^] |
| Dose 1 Efficacy Lag | 14 days | Pfizer/Moderna [^46^] |
| Dose 2 Efficacy Lag | 14 days | Pfizer/Moderna [^46^] |
| Total Number of Doses | 8,000,000 | Experimental Parameter |
| Vaccine Distribution Time Window | Jan 10th - July 17th, 2021 | Experimental Parameter |
| Pre Vaccine Number of Doses | 200,000 | Experimental Parameter/NC Vaccine Distribution |
| Pre Vaccine Distribution Time Window | Dec 13th 2020 - Jan 10th 2021 | Experimental Parameter/NC Vaccine Distribution |
| Population Crossover threshold | 60% | COVID-19 Vaccination Program Interim Playbook for Jurisdictions Operations Annex [^48^] |
| First dose vs second dose prioritization | No prioritization given | Experimental parameter/no specific guidance to prioritize one vs the other |

**Figure A8**: Key Vaccine Model Parameters

6. **Validation**

The model was validated against cases, hospitalizations, and deaths for North Carolina through April 15th, 2021. The primary metrics were deaths and hospitalizations as there is more certainty around these data values. Lab-confirmed cases were multiplied by the lab case multiplier (discussed in Section 2.2) to convert lab-confirmed cases to true cases for validation. On average, the absolute deviation over the course of the simulation is 8%,5.5%, and 4.9% for deaths, hospitalization, and cases respectively over the course of the simulation through April 15th. The absolute deviation on the last day of the validation, April 15th, was 0.9%,6%, and 3% for deaths, hospitalization, and cases, respectively.

Figure A8 shows the model validation of cases, hospitalizations and death by age group, race, and ethnicity, specifically the distribution of each cumulative metric by age group, race, and ethnicity as of April 15th. The model’s estimate of distribution of hospitalizations and deaths by age subgroup differs from reality by at most 4.14% and 6.36%, respectively, tending to overestimate both outcomes for adults (under 65) and underestimate for the older adults (65+) population. The estimate of distribution of hospitalizations and deaths by race and ethnicity differs from reality by at most 3.5% and 2.5%, respectively. The model underestimates the proportion of cases in historically marginalized populations by at most 11.3 % in the Hispanic population. The model also overestimates the proportions of cases in younger populations by 15.8%. This level of accuracy is similar to other published papers^4^.

It is well-documented that the true burden in terms of cases, hospitalizations, and deaths is not the same as reported values. The case burden is known to be higher than the lab-confirmed cases reported^49^. We estimate a lab multiplier as described in the Extended Methodology section to validate cases. The CDC also reports that hospitalizations are underreported^24^. As a reminder, our model estimates cases severe enough for hospitalization, which may not be the same as actual hospitalizations for many reasons including reduced access or care in nursing homes instead of hospitalization. For the Hispanic population, we conjecture the difference in portion of cases is influenced by migrant or seasonal farm and agriculture workers as explained in the Limitations section of the paper.

It is difficult to validate any model during this pandemic. There are many sources of bias in the actual data. Given that we are seeding the model with cases 9 months prior to the date of measurement, we find the model captures a significant portion of what occurs in reality. We also note that the directionality we find between groups is consistent with the events in North Carolina.

| **Subgroup** | **Model Value** | | | **State Value** | | |
| --- | --- | --- | --- | --- | --- | --- |
|  | Cases | Hospitalizations | Deaths | Cases* | Hospitalizations** | Deaths* |
| Age | | | | | | |
| Ages 0-19 | 28% | 3% | 0% | 12.17% | 1.64% | 0.028% |
| Ages 20-64 | 61% | 40% | 10% | 73.52% | 44.14% | 16.33% |
| Ages 65+ | 11% | 58% | 90% | 14.31% | 54.21% | 83.64% |
| Race | | | | | | |
| Black Population | 24.8% | 25.3% | 25.7% | 19.9% | 28.8% | 23.2% |
| Other Population | 7.8% | 7.1% | 6.8% | 17.6 | 8.3% | 8.9% |
| White Population | 67.3% | 67.5% | 67.3% | 62.5% | 62.9% | 67.9% |
| Ethnicity | | | | | | |
| Hispanic Population | 9.6% | 8.9% | 8.5% | 20.2% | 6.6% | 7.8% |
| Non-Hispanic Population | 90.4% | 91.1% | 91.5% | 79.8% | 93.4% | 92.2% |

**Figure A9**: County Level Disease Spread Across 45 Replications

* The North Carolina Department of Health and Human Services (NC DHHS)^29^ reports COVID-19 lab-confirmed cases and deaths using the following age groups: 0-17 years, 18-24 years, 25-49 years, 50-64 years, 65-74 years, and 75+ years. We report estimates for the 0-19 age group using the 0-17 age group.

**NC DHHS^29^ reports COVID-19 related hospitalizations using the following age groups: 0-17 years, 18-19 years, 20-29 years, 30-39 years, 40-49 years, 50-59 years, 60-69 years, 70-79 years, and 80+ years. We estimate hospitalizations corresponding to the 60-64 and 65-69 age groups using a weighted average representative of the North Carolina population.

7. **Stochastic Nature of Simulation and Uncertainty**

Every replication of every scenario begins with a different random seed for the random number generator. No two replications for the same scenario are equivalent. The difference in random seeds leads to differences in the realization of the network structure (household and peer group assignments), race assignment, diabetes assignment, initial seeding of cases and deaths, who wears a mask, who goes to work, who gets vaccinated, all disease interactions, etc.

Multiple statistics are calculated from the output generated by the set of 45 replications, including the average and variance. Goodness of fit tests were conducted on cumulative infections, hospitalizations, and deaths at a state level each day of the simulation and were shown to follow a normal distribution. This allows for parametric statistical tests to be conducted that utilize confidence intervals that are derived from the variance.

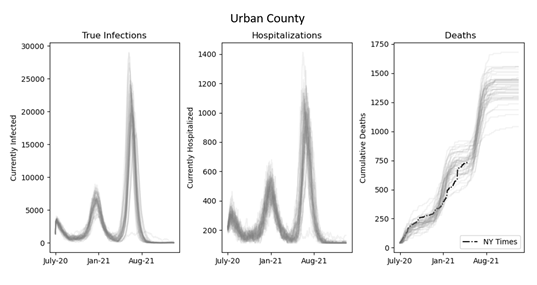

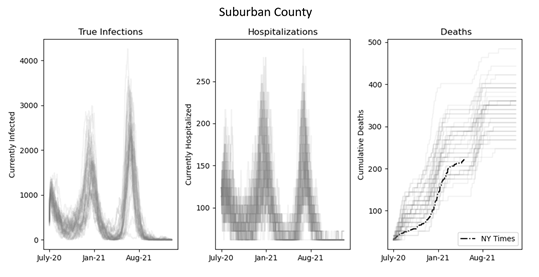

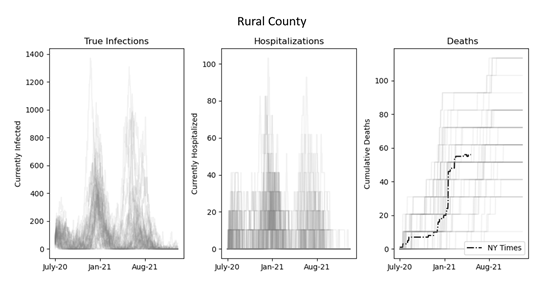

**Figure A10**: County Level Disease Spread Across 45 Replications

Figure A9 demonstrates the variability that arises in the simulation at the county level. There is more variability in the rural county as the aggregated population is smaller. The figure for deaths also displays data summarized in the NY Times, where the value is within the set of replicated values for each of the counties. These figures emphasize that the progression of the disease within the population is stochastic, and that the outcomes are highly dependent on the behaviors and interactions of those who are infected.

8. **Extended Results**

**Infection and Hospitalization Equity Gap**

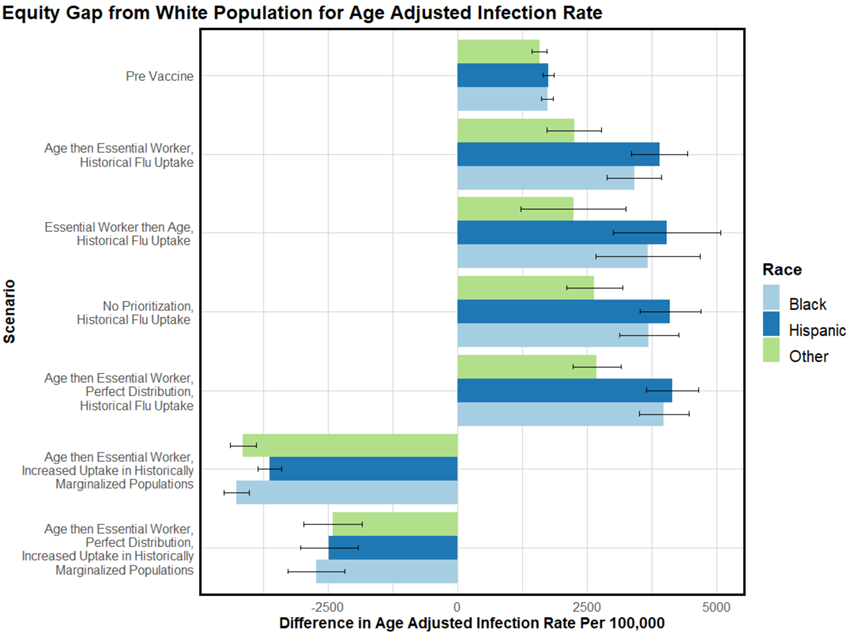

**Figure A11**: Historically Marginalized Populations’ Equity Gap from White Population: Infections

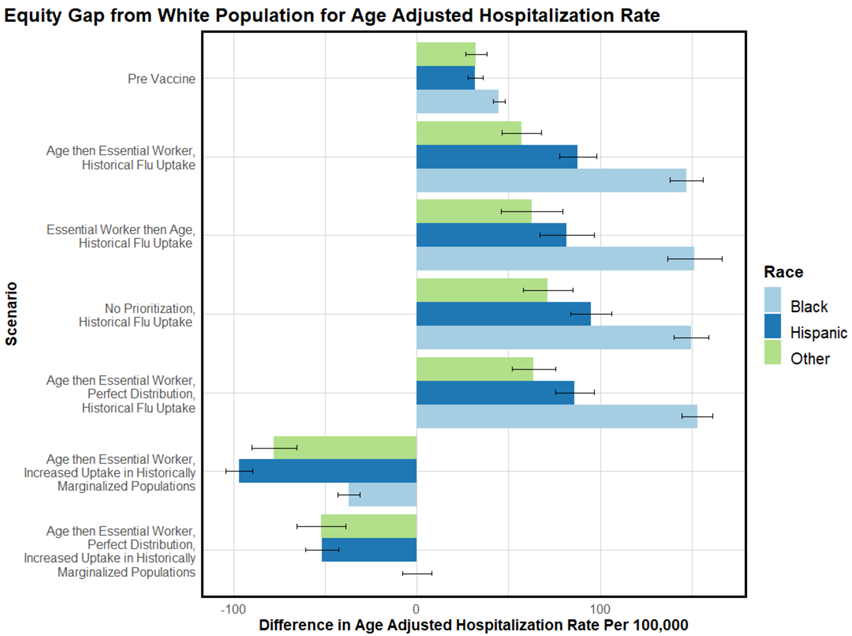

**Figure A12**: Historically Marginalized Populations’ Equity Gap from White Population: Hospitalizations

**Result Tables**

Figures A12/A13/A14 show the cumulative infections, hospitalization, and deaths at the state level and age-adjusted metrics for each subpopulation. The uptake, population 1 and population 2 parameters uniquely determine a scenario. In Figure A12, different combinations of priority populations are tested with historical flu uptake. A subset of these orderings are shown in Figure A13 with increased uptake in HMPs and increased uptake in the entire population. Finally Figure A14 shows these same subset of ordering scenarios with susceptible-only distribution with all three levels of uptake.

| Uptake | Scenario | Population 1 | Population 2 | Cumulative Metric | State | CI | Black (Age Adjusted) | CI | Hispanic (Age Adjusted) | CI | Other (Age Adjusted) | CI | White (Age Adjusted) | CI |
| --- | --- | --- | --- | --- | --- | --- | --- | --- | --- | --- | --- | --- | --- | --- |
| Historical Flu Uptake | Scenario 1 | 65+ | High Risk | Infected | 42.908% | 0.559% | 46.060% | 0.382% | 46.469% | 0.424% | 44.895% | 0.394% | 42.295% | 0.401% |
|  |  |  |  | Hospitalized | 0.798% | 0.010% | 0.843% | 0.008% | 0.784% | 0.009% | 0.757% | 0.014% | 0.690% | 0.006% |
|  |  |  |  | Dead | 0.174% | 0.002% | 0.179% | 0.002% | 0.158% | 0.003% | 0.154% | 0.006% | 0.136% | 0.001% |
|  | Scenario 2 | 65+ | Essential Workers | Infected | 42.908% | 0.528% | 45.829% | 0.363% | 46.311% | 0.387% | 44.666% | 0.356% | 42.412% | 0.381% |
|  |  |  |  | Hospitalized | 0.797% | 0.008% | 0.838% | 0.007% | 0.779% | 0.009% | 0.749% | 0.009% | 0.691% | 0.005% |
|  |  |  |  | Dead | 0.174% | 0.002% | 0.181% | 0.003% | 0.158% | 0.003% | 0.153% | 0.005% | 0.135% | 0.001% |
|  | Scenario 3 | 65+ | Historically Marginalized Populations | Infected | 43.068% | 0.185% | 45.185% | 0.103% | 45.605% | 0.136% | 44.020% | 0.175% | 43.018% | 0.176% |
|  |  |  |  | Hospitalized | 0.804% | 0.003% | 0.835% | 0.006% | 0.770% | 0.007% | 0.743% | 0.010% | 0.703% | 0.004% |
|  |  |  |  | Dead | 0.175% | 0.001% | 0.182% | 0.002% | 0.153% | 0.003% | 0.151% | 0.005% | 0.137% | 0.001% |
|  | Scenario 4 | 65+ | Low Income | Infected | 43.583% | 0.176% | 46.463% | 0.126% | 47.117% | 0.142% | 45.706% | 0.181% | 43.082% | 0.146% |
|  |  |  |  | Hospitalized | 0.809% | 0.004% | 0.850% | 0.005% | 0.797% | 0.008% | 0.766% | 0.010% | 0.701% | 0.003% |
|  |  |  |  | Dead | 0.175% | 0.001% | 0.181% | 0.002% | 0.161% | 0.003% | 0.152% | 0.005% | 0.137% | 0.001% |
|  | Scenario 5 | High Risk | 65+ | Infected | 43.106% | 0.181% | 46.283% | 0.141% | 46.748% | 0.167% | 45.011% | 0.169% | 42.479% | 0.144% |
|  |  |  |  | Hospitalized | 0.800% | 0.003% | 0.846% | 0.005% | 0.785% | 0.008% | 0.748% | 0.011% | 0.691% | 0.003% |
|  |  |  |  | Dead | 0.177% | 0.001% | 0.184% | 0.003% | 0.157% | 0.003% | 0.156% | 0.005% | 0.138% | 0.001% |
|  | Scenario 6 | Essential Workers | 65+ | Infected | 41.093% | 1.029% | 44.140% | 0.708% | 44.505% | 0.744% | 42.704% | 0.716% | 40.463% | 0.721% |
|  |  |  |  | Hospitalized | 0.783% | 0.017% | 0.827% | 0.012% | 0.757% | 0.012% | 0.738% | 0.014% | 0.675% | 0.009% |
|  |  |  |  | Dead | 0.174% | 0.003% | 0.180% | 0.003% | 0.157% | 0.004% | 0.155% | 0.005% | 0.135% | 0.002% |
|  | Scenario 7 | Historically Marginalized Populations | 65+ | Infected | 43.119% | 0.204% | 44.869% | 0.115% | 45.479% | 0.147% | 43.776% | 0.164% | 43.215% | 0.195% |
|  |  |  |  | Hospitalized | 0.810% | 0.004% | 0.824% | 0.005% | 0.765% | 0.008% | 0.745% | 0.010% | 0.714% | 0.003% |
|  |  |  |  | Dead | 0.177% | 0.001% | 0.179% | 0.002% | 0.157% | 0.003% | 0.153% | 0.005% | 0.140% | 0.001% |
|  | Scenario 8 | Low Income | 65+ | Infected | 43.530% | 0.158% | 46.313% | 0.126% | 46.999% | 0.131% | 45.687% | 0.181% | 43.045% | 0.129% |
|  |  |  |  | Hospitalized | 0.813% | 0.003% | 0.846% | 0.005% | 0.803% | 0.008% | 0.761% | 0.011% | 0.706% | 0.003% |
|  |  |  |  | Dead | 0.177% | 0.001% | 0.182% | 0.002% | 0.161% | 0.003% | 0.153% | 0.005% | 0.139% | 0.001% |
|  | Scenario 9 | No Prioritization | | Infected | 42.187% | 0.569% | 45.246% | 0.401% | 45.655% | 0.431% | 44.193% | 0.367% | 41.547% | 0.398% |
|  |  |  |  | Hospitalized | 0.802% | 0.010% | 0.841% | 0.007% | 0.786% | 0.010% | 0.763% | 0.012% | 0.691% | 0.006% |
|  |  |  |  | Dead | 0.178% | 0.002% | 0.184% | 0.002% | 0.162% | 0.003% | 0.160% | 0.005% | 0.139% | 0.001% |

**Figure A13**: Prioritization Ordering Scenarios

| Uptake | Scenario | Population 1 | Population 2 | Cumulative Metric | State | CI | Black (Age Adjusted) | CI | Hispanic (Age Adjusted) | CI | Other (Age Adjusted) | CI | White (Age Adjusted) | CI |
| --- | --- | --- | --- | --- | --- | --- | --- | --- | --- | --- | --- | --- | --- | --- |
| Increased HMP Uptake | Scenario 10 | 65+ | Essential Workers | Infected | 43.630% | 0.207% | 41.532% | 0.165% | 42.172% | 0.152% | 41.655% | 0.184% | 45.792% | 0.174% |
|  |  |  |  | Hospitalized | 0.797% | 0.004% | 0.716% | 0.005% | 0.656% | 0.006% | 0.675% | 0.012% | 0.753% | 0.003% |
|  |  |  |  | Dead | 0.169% | 0.001% | 0.147% | 0.002% | 0.130% | 0.003% | 0.134% | 0.005% | 0.144% | 0.001% |
|  | Scenario 11 | 65+ | Historically Marginalized Populations | Infected | 44.626% | 0.243% | 38.857% | 0.120% | 39.774% | 0.117% | 39.529% | 0.165% | 48.754% | 0.264% |
|  |  |  |  | Hospitalized | 0.811% | 0.004% | 0.670% | 0.005% | 0.627% | 0.007% | 0.636% | 0.009% | 0.795% | 0.004% |
|  |  |  |  | Dead | 0.171% | 0.001% | 0.144% | 0.002% | 0.124% | 0.003% | 0.133% | 0.004% | 0.149% | 0.001% |
|  | Scenario 12 | Essential Workers | 65+ | Infected | 40.644% | 0.678% | 37.686% | 0.323% | 38.458% | 0.337% | 37.850% | 0.331% | 42.979% | 0.544% |
|  |  |  |  | Hospitalized | 0.808% | 0.015% | 0.717% | 0.007% | 0.662% | 0.008% | 0.672% | 0.009% | 0.754% | 0.010% |
|  |  |  |  | Dead | 0.186% | 0.003% | 0.163% | 0.002% | 0.144% | 0.004% | 0.153% | 0.005% | 0.157% | 0.003% |
|  | Scenario 13 | Historically Marginalized Populations | 65+ | Infected | 44.356% | 0.366% | 35.974% | 0.171% | 36.793% | 0.157% | 36.641% | 0.199% | 49.829% | 0.402% |
|  |  |  |  | Hospitalized | 0.836% | 0.007% | 0.644% | 0.006% | 0.597% | 0.007% | 0.614% | 0.011% | 0.838% | 0.007% |
|  |  |  |  | Dead | 0.182% | 0.001% | 0.145% | 0.003% | 0.127% | 0.002% | 0.132% | 0.005% | 0.161% | 0.001% |
| Increased All Populations Uptake | Scenario 14 | 65+ | Essential Workers | Infected | 43.689% | 0.261% | 45.316% | 0.201% | 45.872% | 0.203% | 45.434% | 0.226% | 43.935% | 0.217% |
|  |  |  |  | Hospitalized | 0.774% | 0.005% | 0.788% | 0.007% | 0.731% | 0.008% | 0.745% | 0.012% | 0.689% | 0.004% |
|  |  |  |  | Dead | 0.161% | 0.001% | 0.162% | 0.002% | 0.140% | 0.003% | 0.144% | 0.004% | 0.128% | 0.001% |
|  | Scenario 15 | 65+ | Historically Marginalized Populations | Infected | 45.143% | 0.250% | 40.502% | 0.122% | 41.312% | 0.131% | 40.989% | 0.146% | 48.852% | 0.270% |
|  |  |  |  | Hospitalized | 0.782% | 0.003% | 0.702% | 0.006% | 0.644% | 0.006% | 0.657% | 0.009% | 0.749% | 0.005% |
|  |  |  |  | Dead | 0.160% | 0.001% | 0.150% | 0.002% | 0.128% | 0.002% | 0.136% | 0.005% | 0.133% | 0.001% |
|  | Scenario 16 | Essential Workers | 65+ | Infected | 39.034% | 0.629% | 40.429% | 0.384% | 40.885% | 0.407% | 40.531% | 0.391% | 38.955% | 0.444% |
|  |  |  |  | Hospitalized | 0.800% | 0.014% | 0.795% | 0.008% | 0.737% | 0.010% | 0.733% | 0.013% | 0.701% | 0.008% |
|  |  |  |  | Dead | 0.188% | 0.003% | 0.182% | 0.003% | 0.162% | 0.003% | 0.162% | 0.006% | 0.150% | 0.002% |
|  | Scenario 17 | Historically Marginalized Populations | 65+ | Infected | 45.017% | 0.242% | 37.192% | 0.129% | 37.951% | 0.122% | 37.728% | 0.195% | 50.318% | 0.267% |
|  |  |  |  | Hospitalized | 0.821% | 0.004% | 0.664% | 0.005% | 0.605% | 0.007% | 0.615% | 0.010% | 0.816% | 0.005% |
|  |  |  |  | Dead | 0.174% | 0.001% | 0.147% | 0.002% | 0.126% | 0.003% | 0.133% | 0.005% | 0.152% | 0.001% |

**Figure A14**: Increased Uptake Scenarios

| Uptake | Scenario | Population 1 | Population 2 | Cumulative Metric | State | CI | Black (Age Adjusted) | CI | Hispanic (Age Adjusted) | CI | Other (Age Adjusted) | CI | White (Age Adjusted) | CI |
| --- | --- | --- | --- | --- | --- | --- | --- | --- | --- | --- | --- | --- | --- | --- |
| Perfect Distribution | Scenario 18 | 65+ | Essential Workers | Infected | 40.910% | 0.488% | 44.202% | 0.342% | 44.368% | 0.373% | 42.905% | 0.316% | 40.216% | 0.341% |
|  |  |  |  | Hospitalized | 0.761% | 0.008% | 0.809% | 0.007% | 0.742% | 0.009% | 0.720% | 0.011% | 0.656% | 0.005% |
|  |  |  |  | Dead | 0.167% | 0.001% | 0.174% | 0.002% | 0.156% | 0.004% | 0.148% | 0.005% | 0.129% | 0.001% |
|  | Scenario 19 | 65+ | Historically Marginalized Populations | Infected | 40.968% | 0.474% | 44.022% | 0.341% | 44.333% | 0.349% | 42.601% | 0.327% | 40.390% | 0.333% |
|  |  |  |  | Hospitalized | 0.762% | 0.008% | 0.808% | 0.007% | 0.742% | 0.008% | 0.714% | 0.010% | 0.658% | 0.005% |
|  |  |  |  | Dead | 0.167% | 0.002% | 0.174% | 0.002% | 0.152% | 0.003% | 0.151% | 0.005% | 0.130% | 0.001% |
|  | Scenario 20 | Essential Workers | 65+ | Infected | 36.855% | 1.507% | 40.026% | 1.041% | 40.035% | 1.114% | 38.830% | 0.987% | 36.055% | 1.048% |
|  |  |  |  | Hospitalized | 0.711% | 0.025% | 0.758% | 0.015% | 0.690% | 0.017% | 0.677% | 0.016% | 0.610% | 0.014% |
|  |  |  |  | Dead | 0.160% | 0.004% | 0.167% | 0.004% | 0.143% | 0.005% | 0.145% | 0.005% | 0.125% | 0.003% |
|  | Scenario 21 | Historically Marginalized Populations | 65+ | Infected | 40.611% | 0.752% | 43.327% | 0.513% | 43.724% | 0.544% | 41.870% | 0.512% | 40.168% | 0.532% |
|  |  |  |  | Hospitalized | 0.764% | 0.013% | 0.796% | 0.009% | 0.735% | 0.011% | 0.692% | 0.010% | 0.666% | 0.007% |
|  |  |  |  | Dead | 0.171% | 0.003% | 0.174% | 0.003% | 0.150% | 0.003% | 0.145% | 0.005% | 0.135% | 0.002% |
| Perfect Distribution Increased HMP Uptake | Scenario 22 | 65+ | Essential Workers | Infected | 35.388% | 0.535% | 34.142% | 0.281% | 34.393% | 0.293% | 34.469% | 0.310% | 36.869% | 0.469% |
|  |  |  |  | Hospitalized | 0.666% | 0.007% | 0.617% | 0.005% | 0.565% | 0.006% | 0.565% | 0.012% | 0.617% | 0.006% |
|  |  |  |  | Dead | 0.149% | 0.001% | 0.136% | 0.002% | 0.115% | 0.003% | 0.121% | 0.005% | 0.125% | 0.002% |
|  | Scenario 23 | 65+ | Historically Marginalized Populations | Infected | 36.080% | 0.489% | 33.422% | 0.194% | 33.655% | 0.235% | 33.937% | 0.260% | 38.351% | 0.490% |
|  |  |  |  | Hospitalized | 0.674% | 0.007% | 0.600% | 0.005% | 0.540% | 0.007% | 0.559% | 0.012% | 0.636% | 0.007% |
|  |  |  |  | Dead | 0.149% | 0.001% | 0.132% | 0.002% | 0.111% | 0.003% | 0.120% | 0.004% | 0.126% | 0.001% |
|  | Scenario 24 | Essential Workers | 65+ | Infected | 31.522% | 1.021% | 31.072% | 0.484% | 30.981% | 0.545% | 31.049% | 0.555% | 32.439% | 0.861% |
|  |  |  |  | Hospitalized | 0.624% | 0.016% | 0.591% | 0.006% | 0.533% | 0.007% | 0.544% | 0.009% | 0.567% | 0.011% |
|  |  |  |  | Dead | 0.144% | 0.003% | 0.135% | 0.002% | 0.115% | 0.003% | 0.121% | 0.004% | 0.119% | 0.002% |
|  | Scenario 25 | Historically Marginalized Populations | 65+ | Infected | 36.317% | 0.348% | 32.448% | 0.181% | 32.773% | 0.184% | 32.937% | 0.217% | 39.206% | 0.367% |
|  |  |  |  | Hospitalized | 0.693% | 0.006% | 0.599% | 0.005% | 0.535% | 0.006% | 0.554% | 0.010% | 0.662% | 0.005% |
|  |  |  |  | Dead | 0.155% | 0.001% | 0.135% | 0.002% | 0.117% | 0.003% | 0.117% | 0.004% | 0.133% | 0.001% |
| Perfect Distribution Increased All Populations Uptake | Scenario 26 | 65+ | Essential Workers | Infected | 35.226% | 0.484% | 37.303% | 0.309% | 37.487% | 0.339% | 37.787% | 0.313% | 34.972% | 0.389% |
|  |  |  |  | Hospitalized | 0.638% | 0.005% | 0.664% | 0.005% | 0.598% | 0.008% | 0.626% | 0.010% | 0.559% | 0.005% |
|  |  |  |  | Dead | 0.139% | 0.001% | 0.145% | 0.002% | 0.119% | 0.002% | 0.129% | 0.004% | 0.109% | 0.001% |
|  | Scenario 27 | 65+ | Historically Marginalized Populations | Infected | 36.350% | 0.602% | 34.770% | 0.263% | 35.157% | 0.303% | 35.250% | 0.339% | 38.128% | 0.567% |
|  |  |  |  | Hospitalized | 0.655% | 0.007% | 0.627% | 0.005% | 0.571% | 0.007% | 0.586% | 0.011% | 0.603% | 0.007% |
|  |  |  |  | Dead | 0.141% | 0.001% | 0.140% | 0.002% | 0.117% | 0.002% | 0.125% | 0.005% | 0.114% | 0.001% |
|  | Scenario 28 | Essential Workers | 65+ | Infected | 30.709% | 0.763% | 32.870% | 0.481% | 32.663% | 0.532% | 32.983% | 0.554% | 30.173% | 0.556% |
|  |  |  |  | Hospitalized | 0.615% | 0.013% | 0.639% | 0.008% | 0.573% | 0.009% | 0.598% | 0.012% | 0.530% | 0.007% |
|  |  |  |  | Dead | 0.144% | 0.002% | 0.146% | 0.003% | 0.124% | 0.003% | 0.132% | 0.005% | 0.114% | 0.002% |
|  | Scenario 29 | Historically Marginalized Populations | 65+ | Infected | 36.120% | 0.618% | 32.713% | 0.245% | 33.121% | 0.288% | 33.371% | 0.285% | 38.824% | 0.621% |
|  |  |  |  | Hospitalized | 0.668% | 0.009% | 0.599% | 0.006% | 0.545% | 0.006% | 0.559% | 0.008% | 0.631% | 0.008% |
|  |  |  |  | Dead | 0.146% | 0.001% | 0.136% | 0.002% | 0.114% | 0.003% | 0.122% | 0.005% | 0.122% | 0.001% |

**Figure A15**: Increased Uptake and Susceptible-only Distribution Scenarios

9. **Realization of Vaccine Distribution**

The total number of vaccines distributed and accepted by each population over time is tracked during the simulation. Figure A15 shows the proportion of older adults (65+), and proportion of all agents who have received their first/second dose. Figure A16 shows the number of essential workers who have received the first and second doses. Both figures show the baseline scenario of age then essential workers being prioritized with historical flu uptake levels.

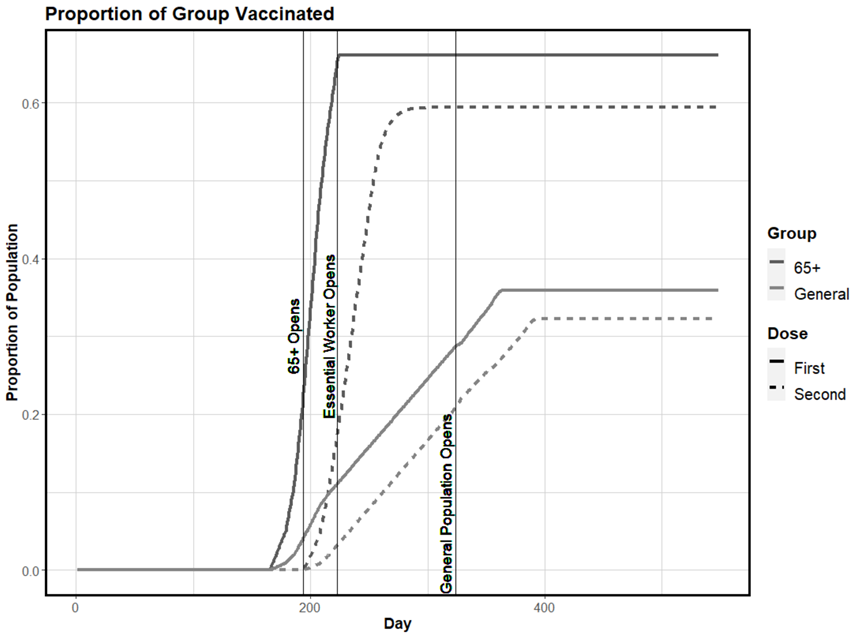

**Figure A16**: Baseline Proportion of Older Adult and Total Population Receiving First and Second Dose

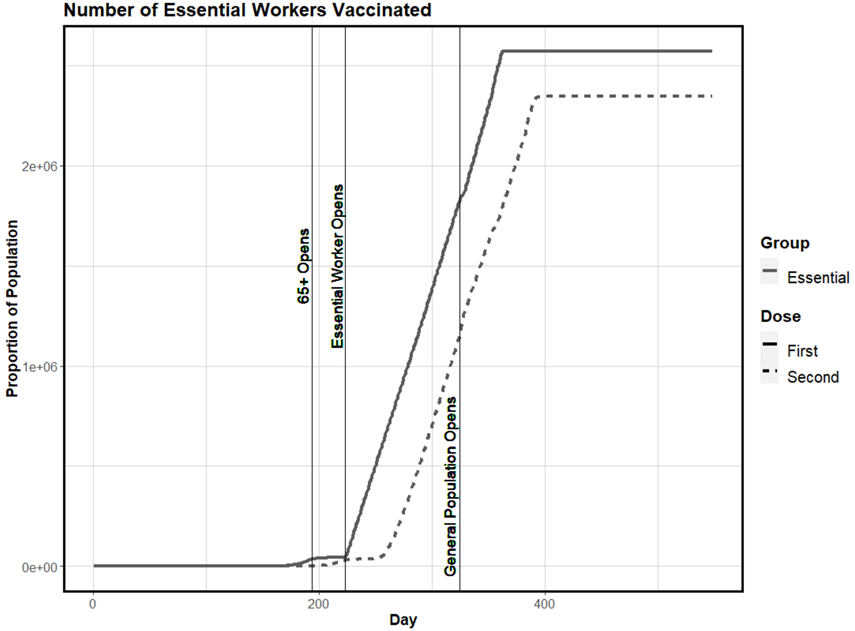

**Figure A17**: Baseline Number of Essential Workers Receiving First and Second Dose

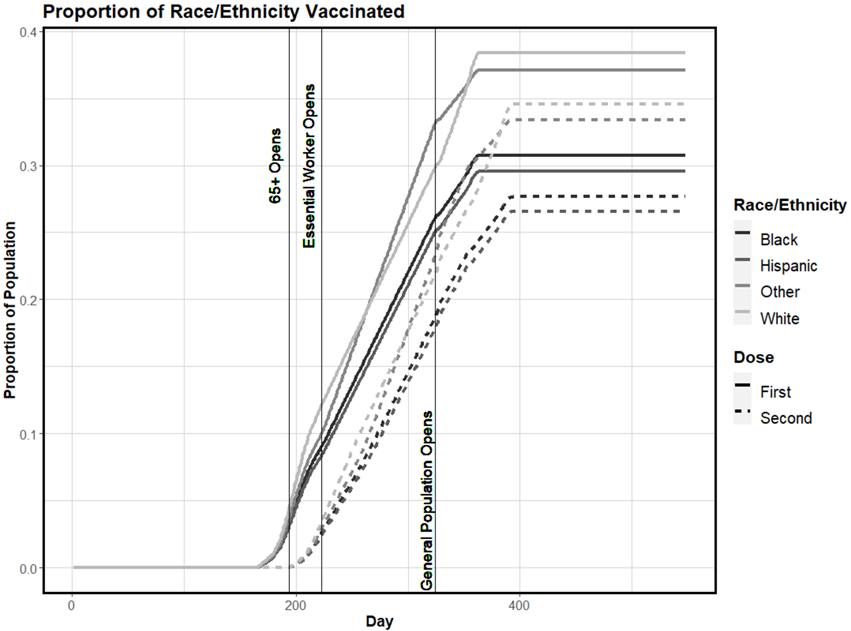

**Figure A18**: Baseline Proportion of Total Population Receiving First and Second Dose by Race/Ethnicity

5.	American Community Survey 5-year Estimates 2017 [database on the Internet] [cited January 2021]. Available from: data.census.gov.

6.	Census Summary File 1 [database on the Internet]2010. Available from: data.census.gov.

12.	COVID-19 Community Mobility Reports [database on the Internet]. Available from: <https://www.google.com/covid19/mobility/>.

13.	Statista. Race and ethnicity of U.S. households in 2015, by size. Statista: Statista Research Department; 2021 [cited 2021 July]; Available from: <https://www.statista.com/statistics/638956/race-and-ethnicity-of-us-households-by-size/>.

14.	Schaeffer K. The most common age among whites in U.S. is 58 – more than double that of racial and ethnic minorities. Pew Research Center2019 [cited 2021 July 5]; Available from: <https://www.pewresearch.org/fact-tank/2019/07/30/most-common-age-among-us-racial-ethnic-groups/>.

15.	CDC. COVID-19 Vaccination Program Interim Playbook for Jurisdictions Operations Annex. Centers for Disease Control and Prevention2021 January 2021.

16.	Fujimoto AB, Yildirim I, Keskinocak P. Significance of SARS-CoV-2 Specific Antibody Testing during COVID-19 Vaccine Allocation. Vaccine. 2021 2021/06/26/.

17.	Delphi Epidata API [database on the Internet]2021. Available from: <https://github.com/cmu-delphi/delphi-epidata>.

18.	Porter F. Governor Cooper Signs School Reopening Bill into Law. NC Governor Roy Cooper2021.

19.	CDC. COVID Data Tracker. Centers for Disease Control and Prevention2020-2021; Available from: <https://covid.cdc.gov/covid-data-tracker/#mobility>.

20.	MacMillan D. Following New CDC Guidance on Face Coverings, Governor Cooper Lifts Many COVID-19 Restrictions. NC Governor Roy Cooper2021.

32.	Ferguson NM LD, Nedjati-Gilani G, Imai N, Ainslie K, Baguelin M, Bhatia S, et al. Report 9: Impact of non-pharmaceutical interventions (NPIs) to

reduce COVID-19 mortality and healthcare demand2020.

40.	Team CC-R. Severe Outcomes Among Patients with Coronavirus Disease 2019 (COVID-19) — United States, February 12–March 16, 20202020.

41.	CDC Case Surveillance Team DAaMTF. COVID-19 Case Surveillance Restricted Access Detailed Data. CDC2020-2021.

42.	Gregory JM, Slaughter JC, Duffus SH, Smith TJ, LeStourgeon LM, Jaser SS, et al. COVID-19 Severity Is Tripled in the Diabetes Community: A Prospective Analysis of the Pandemic’s Impact in Type 1 and Type 2 Diabetes. Diabetes Care. 2021;44(2):526.

43.	Li R, Pei S, Chen B, Song Y, Zhang T, Yang W, et al. Substantial undocumented infection facilitates the rapid dissemination of novel coronavirus (SARS-CoV-2). Science. 2020;368(6490):489-93.

44.	Organization WH. Report of the WHO-China Joint Mission on Coronavirus Disease 2019 (COVID-19). World Health Organization2020 February 2020.

45.	Walker PGT, Whittaker C, Watson OJ, Baguelin M, Winskill P, Hamlet A, et al. The impact of COVID-19 and strategies for mitigation and suppression in low- and middle-income countries. Science. 2020;369(6502):413-22.

46.	CDC. Science Brief: COVID-19 Vaccines and Vaccination. Centers for Disease Control and Prevention2021; Available from: <https://www.cdc.gov/coronavirus/2019-ncov/science/science-briefs/fully-vaccinated-people.html>.

47.	Kriss JL, Reynolds LE, Wang A, Stokley S, Cole MM, Harris LQ, et al. COVID-19 Vaccine Second-Dose Completion and Interval Between First and Second Doses Among Vaccinated Persons - United States, December 14, 2020-February 14,20212021.

48.	CDC. COVID-19 Vaccination Program Interim Playbook for Jurisdictions Operations Annex. Centers for Disease Control and Prevention2021 January 2021.

49.	MacDougall R. NIH study suggests COVID-19 prevalence far exceeded early pandemic cases2021: Available from: <https://www.nih.gov/news-events/news-releases/nih-study-suggests-covid-19-prevalence-far-exceeded-early-pandemic-cases>.
